# Mammographic density, pathogenic breast cancer susceptibility gene variants and breast cancer risk

**DOI:** 10.1101/2025.04.17.25325994

**Authors:** Xiaomeng Zhang, Mikael Eriksson, Nasim Mavaddat, Joe Dennis, Susan M. Astley, Marike Gabrielson, Graham G. Giles, Steven N. Hart, David J. Hunter, Loic Le Marchand, Michael Lush, Kyriaki Michailidou, Christopher G. Scott, Qin Wang, Sacha J. Howell, Marc Naven, Antonis C. Antoniou, Kristan J. Aronson, Manjeet K. Bolla, Jose E. Castelao, Fergus J. Couch, Kamila Czene, Alison M. Dunning, D. Gareth Evans, Manuela Gago-Dominguez, Montserrat García-Closas, Christopher A. Haiman, Roger L. Milne, Paul D.P. Pharoah, Melissa C. Southey, Jennifer Stone, Rachel A. Murphy, Amy Berrington de Gonzalez, Celine M. Vachon, Per Hall, Douglas F Easton

**Affiliations:** Centre for Cancer Genetic Epidemiology, Department of Public Health and Primary Care. University of Cambridge. Cambridge: UK; CB1 8RN; Department of Medical Epidemiology and Biostatistics. Karolinska Institutet. Stockholm: Sweden; 171 65; Division of Informatics, Imaging and Data Sciences, Faculty of Biology, Medicine and Health. University of Manchester, Manchester Academic Health Science Centre. Manchester: UK; M13 9PT; Cancer Epidemiology Division. Cancer Council Victoria. Melbourne, Victoria: Australia; 3004; Centre for Epidemiology and Biostatistics, Melbourne School of Population and Global Health. The University of Melbourne. Melbourne, Victoria: Australia; 3010; Precision Medicine, School of Clinical Sciences at Monash Health. Monash University. Clayton, Victoria: Australia; 3168; Department of Health Sciences Research. Mayo Clinic. Rochester, MN: USA; 55905; Department of Laboratory Medicine and Pathology. Mayo Clinic. Rochester, MN: USA; 55905; Nuffield Department of Population Health. University of Oxford. Oxford: UK; OX3 7LF; Department of Epidemiology. Harvard TH Chan School of Public Health. Boston, MA: USA; 02115; Epidemiology Program. University of Hawaii Cancer Center. Honolulu, HI: USA; 96813; Biostatistics Unit. The Cyprus Institute of Neurology and Genetics. Nicosia: Cyprus; 2371; Department of Quantitative Health Sciences, Division of Clinical Trials and Biostatistics. Mayo Clinic. Rochester, MN: USA; 55905; Division of Cancer Sciences. University of Manchester. Manchester: UK; M20 4GJ; Department of Public Health Sciences and Sinclair Cancer Research Institute. Queen’s University. Kingston, ON: Canada; K7L 3N6; Oncology and Genetics Group. Galicia Sur Health Research Institute (IIS Galicia Sur) SERGAS-UVIGO. Vigo: Spain; 36312; Centre for Cancer Genetic Epidemiology, Department of Oncology. University of Cambridge. Cambridge: UK; CB1 8RN; Division of Evolution and Genomic Sciences, School of Biological Sciences, Faculty of Biology, Medicine and Health. University of Manchester, Manchester Academic Health Science Centre. Manchester: UK; M13 9WL; North West Genomics Laboratory Hub, Manchester Centre for Genomic Medicine. St Mary’s Hospital, Manchester University NHS Foundation Trust, Manchester Academic Health Science Centre. Manchester: UK; M13 9WL; Cancer Genetics and Epidemiology Group, Genomic Medicine Group. Fundación Instituto de Investigación Sanitaria de Santiago de Compostela (FIDIS), Complejo Hospitalario Universitario de Santiago, SERGAS. Santiago de Compostela: Spain; 15706; Division of Genetics and Epidemiology. The Institute of Cancer Research. London: UK; SM2 5NG; Center for Genetic Epidemiology, Department of Population and Public Health Sciences, Keck School of Medicine. University of Southern California. Los Angeles, CA: USA; 90033; Department of Computational Biomedicine. Cedars-Sinai Medical Center. West Hollywood, CA: USA; 90069; Department of Clinical Pathology. The University of Melbourne. Melbourne, Victoria: Australia; 3010; Genetic Epidemiology Group, School of Population and Global Health. University of Western Australia. Perth, Western Australia: Australia; 6000; School of Population and Public Health. University of British Columbia. Vancouver, BC: Canada; V6T 1Z4; Cancer Control Research. BC Cancer Agency. Vancouver, BC: Canada; V5Z 1L3; Department of Quantitative Health Sciences, Division of Epidemiology. Mayo Clinic. Rochester, MN: USA; 55905; Department of Oncology. Södersjukhuset. Stockholm: Sweden; 118 83

## Abstract

**Importance:** Mammographic density (MD) and pathogenic variants (PVs) in breast cancer susceptibility genes are major determinants of breast cancer risk, but their association and joint effects on breast cancer risk are unclear.

**Objective:** To investigate the association between the presence or absence of PVs in breast cancer susceptibility genes and MD measures, and their joint effects on breast cancer risk in an observational study; and to evaluate causality using Mendelian randomisation (MR) analyses.

**Design:** Case-control analyses using data from the Breast Cancer Association Consortium (1991-2016). Sequencing and genotyping took place between 2009 and 2021.

**Setting:** Multicenter

**Participants:** A total of 6,809 cases and 18,189 controls were included, from 15 studies, comprising women aged 19 to 92 years with mammograms taken at least one year before diagnosis.

**Exposure:** MD measures, including dense area (DA), non-dense area (NDA), percentage density (PD) and absolute difference in PD between left and right breasts (ADPD), and PVs in *ATM, BARD1, BRCA1, BRCA2, CHEK2, PALB2, RAD51C* and *RAD51D*.

**Main outcomes and measures:** Breast cancer risk overall, by oestrogen receptor expression-defined subtypes, and among *BRCA1* and *BRCA2* PV carriers.

**Results:** No association was found between the overall burden of PVs and any MD measure. There was some evidence for a negative interaction between the burden of PVs in the eight genes and PD (OR=0.79,95%CI_int_=0.62,1.00, P_LRT_=0.047). This appears to be largely driven by a positive interaction with NDA. MR analyses indicated attenuated effects for *BRCA1* (for PD, OR per standard deviation =1.02(95%CI:0.78,1.34) but not *BRCA2* PV carriers (1.54,95%CI=1.08,2.24)).

**Conclusions and Relevance:** There was no evidence of association between PVs in breast cancer susceptibility genes and MD measures, but some suggestion that the association between MD and breast cancer risk may be weaker in PV carriers. Replication of these findings in further large datasets is required.

## Introduction

Breast cancer is the most common cancer in women worldwide. The risk of breast cancer is influenced by both lifestyle and genetic factors, including common variants identified through genome-wide association studies (GWAS) and rare coding variants in susceptibility genes conferring moderate to high risks of the disease.^1, 2^ Aside from genetic risk variants, one of the strongest risk factors is breast density. Mammographic density (MD), assessed on the mammogram, represents the relative proportion of fibroglandular to fatty tissue in the breast: high density is associated with an increased risk of the disease. MD is highly heritable, with > 60% of the variance estimated to be due to genetic factors in twin studies. ^3^ More than 30 genetic loci associated with MD have been identified.^4-6^

Previous analyses indicate that polygenic risk scores for breast cancer are weakly correlated with MD and are independently predictive of breast cancer risk. ^7, 8^ However, it is less clear whether this relationship holds for rarer, moderate/high-penetrance variants in susceptibility genes. While analyses of MD have been conducted in studies of *BRCA1* and *BRCA2* pathogenic variant (PV) carriers, the results from these studies have been inconclusive, with some showing strong evidence of an association and others showing no association. ^9-11^

Here, we investigate the associations between PVs in breast cancer susceptibility genes and MD and the evidence for interaction between these PVs and MD in determining breast cancer risk. As a complementary approach, we assessed the association between genetically predicted MD measures and breast cancer risk using Mendelian Randomisation (MR) analyses.

## Methods

### Study participants

MD measures were collected from seven studies involving participants sequenced through the BRIDGES project within the Breast Cancer Association Consortium (BCAC), comprising 3,133 breast cancer cases and 9,451 controls. All individuals were of European ancestry (Table S1). An additional 3,676 cases and 8,738 controls from 14 BCAC studies with *CHEK2* c.1100delC array genotyping and MD data were included in analyses of *CHEK2* PVs. ^12-14^ All studies were approved by the relevant ethical review boards, and all participants included in these studies provided appropriate consent.

### Mammographic density

MD parameters were measured using STRATUS^15^ for the KARMA study, and Cumulus6^16^ for all other studies (Table S1). We utilised four MD parameters: dense area (cm^2^; DA); percent density (PD), non-dense area (cm^2^; NDA), and the absolute difference in PD between left- and right-side breast (ADPD). All the MD was measured by either mediolateral or craniocaudal views by studies. For controls, we used the average MD of both breasts. For cases, we preferentially included mammograms more than one year prior to diagnosis. Where only mammograms at diagnosis were available, we included measurements from the breast contralateral to the tumour. We excluded cases without information on the timing of the mammograms, cases with mammograms within 1 year of diagnosis lacking tumour side information or with (synchronous) bilateral cancer, and individuals without BMI data at the time of the mammogram or recruitment.

### Genetic data

The BRIDGES project performed targeted sequencing of 34 putative breast cancer susceptibility genes in 60,466 female breast cancer cases and 53,461 controls from BCAC. Detailed methods are provided elsewhere.^1^ Analyses focused on the burden of likely pathogenic protein truncating variants (PVs) in eight genes (*ATM, BARD1, BRCA1, BRCA2, CHEK2, PALB2, RAD51C* and *RAD51D;* supplementary notes, Table S2). *BRCA1, BRCA2* and *PALB2* were considered “high-risk” genes and the remaining genes “moderate-risk”. Details of the iCOGS and OncoArray genotyping have been provided elsewhere. ^12-14^

### Statistical methods

Each of the four MD measures was square-root transformed to provide a distribution that was approximately Gaussian. The associations between the burden of PVs in breast cancer susceptibility genes and transformed MD measures were assessed using linear regression, adjusting for study and age and BMI at mammogram as covariates. Subgroup analyses were performed based on the MD measurement methods (Cumulus6 or STRATUS), menopausal status, breast cancer status and estrogen-receptor (ER) status. Sensitivity analyses were performed by further adjusting for menopausal status, family history of breast cancer, parity, age at menarche, age at first childbirth, and current use of HRT.

To assess the combined effect of gene variants and MD on breast cancer risk, we restricted the analysis to genes with at least 10 PV carriers. First, we obtained residuals of the MD measures by fitting the square root of those measures in a generalised linear regression model, respectively, adjusting for age and 1/BMI at mammogram and study.^17^ We then estimated the association between residual of the MD measurements, gene variant burden and breast cancer in a logistic regression model adjusted for age, BMI and study. Departures from a multiplicative model assumption were assessed by fitting an MD × gene burden interaction term and performing likelihood ratio tests (LRT). Subgroup analyses were performed based on the MD measurement method, menopausal status and ER status.

Association and interaction analyses for *CHEK2* c.1100delC and MD, using the iCOGS and OncoArray data, were conducted in the same way, additionally adjusting for the array. The results were then combined with the *CHEK2* PV results from the BRIDGES dataset using a fixed-effect meta-analysis.

### Mendelian Randomisation

To further investigate the association between MD and breast cancer risk among *BRCA1* and *BRCA2* PV gene variant carriers, two-sample MR analyses were performed using summary statistics for breast cancer risk among *BRCA1* and *BRCA2* PV carriers from the Consortium of Investigators of Modifiers of BRCA1/2 (CIMBA), ^18, 19^ and summary statistics from two MD GWAS as instrumental variables (supplementary notes). ^4, 6^ For comparison, we also conducted MR analyses for overall breast cancer and breast cancer subtypes, using summary statistics from GWAS carried out by the BCAC.^12, 20^ We tested the association between SNPs-represented MD measures and breast cancer and its subtypes using the weighted median ^21^ and inverse variance-weighted method.^22^ Cochran’s Q statistic was used to investigate heterogeneity across SNP effects. A non-zero intercept of MR-Egger (at p<0.05) was considered indicative of directional pleiotropy or a violation of the InSIDE assumption. ^23^ Additionally, MR-PRESSO was applied to identify horizontal pleiotropic outliers. ^24^

Analyses were conducted using R, version 4.2.2. Packages ‘MendelianRandomization’, ‘MRPRESSO’ and ‘TwoSampleMR’ were used. Statistical significance was defined as P<0.05.

## Results

### Study characteristics

We analysed data on 3,133 cases and 9,451 controls who had both assessable MD measures and sequencing data on the eight breast cancer susceptibility genes (Table 1). The mean age at mammogram was 57.9 (SD=9.7) years in cases and 59.3 (SD=8.9) in controls. The majority of women were post-menopausal. In the array data, similar participant characteristics were observed, with the exception that controls were younger than cases. (Table S3).

**Table 1.** Characteristics of study participants.

| Characteristics | Controls (N = 9451) | Cases (N = 3133) | P |
| --- | --- | --- | --- |
| Age, years, mean (SD) | 59.33 (8.86) | 57.95 (9.73) | 2.42E-12 |
| BMI, mean (SD) | 26.06 (4.64) | 26.05 (4.49) | 0.669 |
| Postmenopausal, N (%) | 6933/8700 (79.69) | 2181/2498 (87.31) | 8.22E-18 |
| Family history of breast cancer, N (%) |  |  |  |
| With first-degree Family history | 1242/7295 (17.03) | 567/1879 (30.18) | 3.40E-37 |
| First-degree relative age < 50 years | 41/7295 (0.56) | 41/1879 (2.18) |  |
| First-degree relative age >= 50 years | 213/7295 (2.92) | 152/1879 (8.09) |  |
| Parity, N (%) |  |  |  |
| Nulliparous, N (%) | 1100/9137 (12.04) | 423/2950 (14.34) | ref |
| 1 child, N (%) | 1283/9137 (14.04) | 482/2950 (16.34) | 0.796 |
| 2 children, N (%) | 4165/9137 (45.58) | 1253/2950 (42.47) | 2.07E-04 |
| >2 children, N (%) | 2903/9137 (31.77) | 975/2950 (33.05) | 0.001 |
| Age at menarche, years, mean (SD) | 13.1 (1.52) | 13.16 (1.51) | 0.081 |
| Age at first childbirth, years, mean (SD) | 25.54 (4.94) | 25.88 (5) | 0.002 |
| Current use of HRT, N (%) | 687/8210 (8.37) | 505/2745 (18.4) | 4.20E-48 |
| CUMULUS, N (%) | 4153/9451 (43.94) | 1792/3133 (57.2) | 7.71E-38 |
| Percentage density, mean (SD) | 19.60 (17.45) | 24.04 (19.3) | 8.01E-30 |
| ADPD, mean (SD) | 4.33 (5.16) | 5.93 (8.78) | 3.76E-04 |
| Dense area, mean (SD) | 22.67 (22.14) | 31.25 (26.99) | 8.53E-48 |
| Non-dense area, mean (SD) | 139.87 (65.07) | 126.42 (61.4) | 3.20E-21 |

The P-value was derived from either a 2-sample t-test for continuous variables or a chi-squared test for categorical variables; ADPD: The absolute difference in the percentage density between left- and right-side breast.

All the MD measures were associated with breast cancer status (Table S4) with no significant heterogeneity across studies (Table S5). Among the four MD measures, PD had the strongest association with breast cancer risk, with an estimated 1.47-fold increased risk per SD of the residual of PD (95%CI=1.41,1.54; P=1.45×10^-63^) in BRIDGES and an estimated 1.46-fold increase in the iCOGS and OncoArray data (95%CI=1.39,1.52; P=4.54 ×10^-64^). In the subgroup of individuals in BRIDGES, the burden of PVs in *ATM, BRCA1, BRCA2, CHEK2* and *RAD51C* was associated with an increased risk of breast cancer (Table S6).

### Associations between PVs and MD measures

PV carrier status for any single gene was not associated with any MD measure. There was, however, some evidence that overall PV burden was associated with higher NDA and lower PD in cases (NDA: beta=0.34; P=0.103; PD: beta=-0.39; P=0.005; Figure 1, Table S7) and of the opposite effect in controls (NDA: beta=-0.34; P=0.056, P_int_=0.084; PD: beta=0.15; P = 0.244; P_int_ =0.046; Table S7, Figure 1-2).

**Figure 1.**
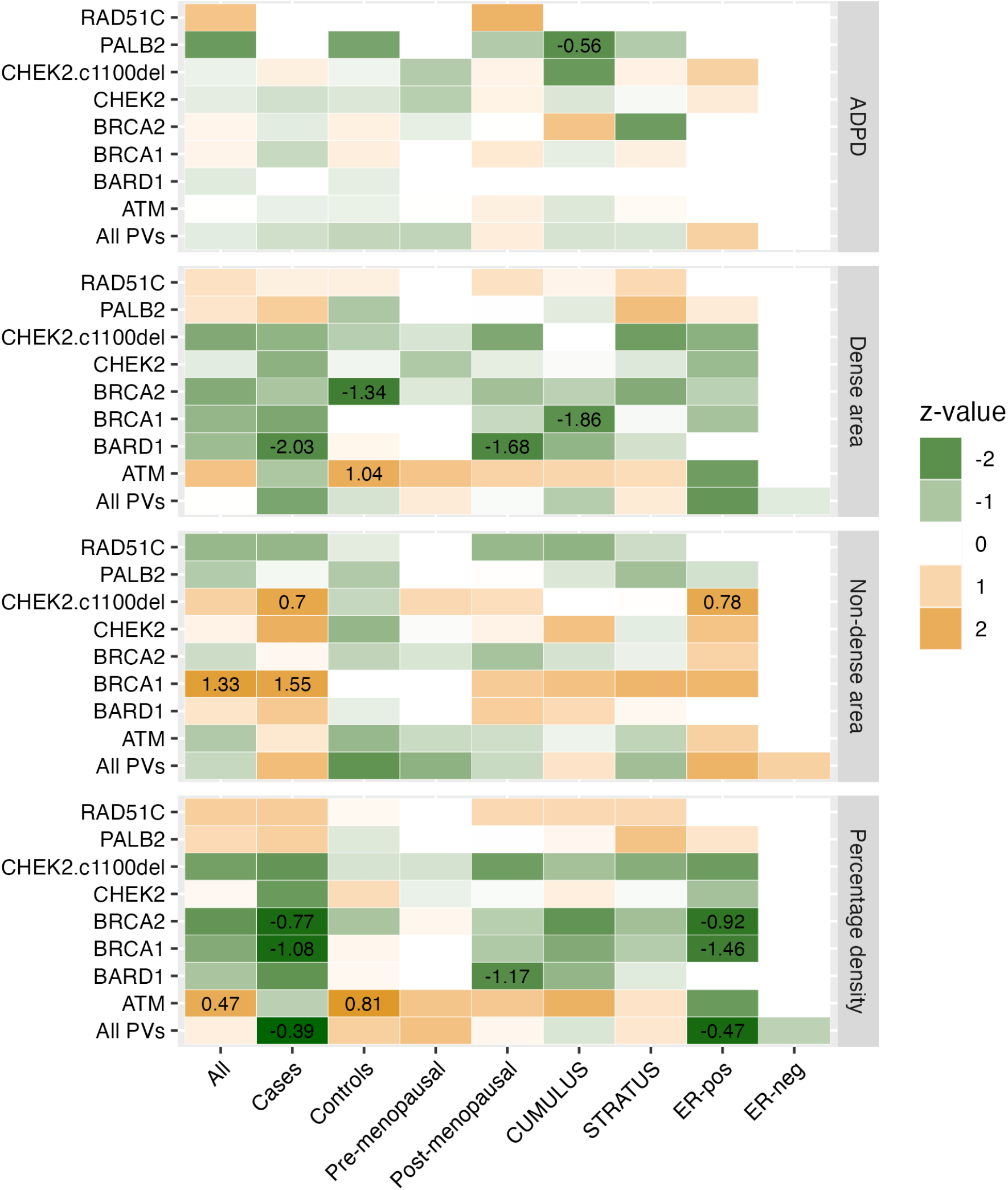
Heatmap of associations between measures of mammographic density and burden of pathogenic variants (PVs) in susceptibility genes. ADPD: absolute difference in PD between left- and right-side breast. The x-axis is the results of overall and subgroup analyses. The y-axis is the breast cancer susceptibility gene. The z-value is two-tailed and estimated by a generalised linear regression model. All labelled associations have a P < 0.05. The labelled number is the estimated beta. The results of *CHEK2*.*c1100del w*ere after a meta-analysis of BRIDGES and array data. All PVs: carrying any PVs across the genes.

**Figure 2.**
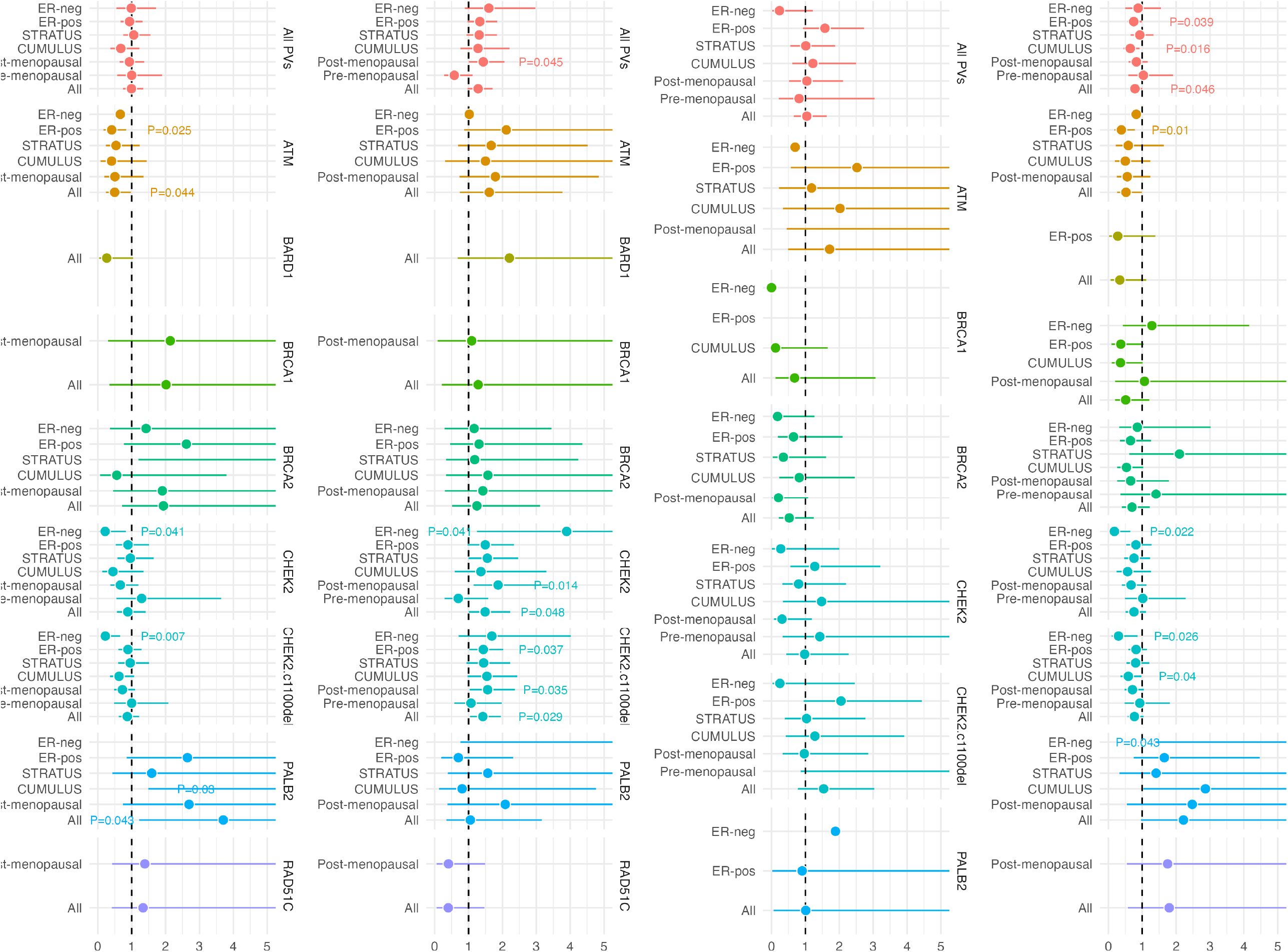
Forest plot of interaction analysis between mammographic density and burden of pathogenic variants (PVs) in relation to breast cancer risk. ADPD: absolute difference in PD between left- and right-side breast. The x-axis is the estimated odds ratio and the corresponding 95% confidence interval. All labelled interactions have a P < 0.05 and at least 10 carriers. The labelled number is the P-value for the interaction term. All PVs: carrying any PVs across the genes.

For individual genes, the strongest association between PV status and MD measures was for *BRCA1*: *BRCA1* PVs were associated with higher NDA (beta=1.33, P=0.016, Table S7). No significant heterogeneity in the effect size was observed by MD measurement method, menopausal status, or ER status. In the meta-analysis of the BRIDGES and array data, there was no evidence of an association between *CHEK2* c.1100delC status and any MD measure (Figure 1, Table S8). Adjusting for additional covariates did not materially affect the results (Table S9-10).

### Combined effect of PVs, MD and breast cancer risk

We next conducted logistic regression analyses with breast cancer as the outcome, and PV carrier status and MD measures were included in the same model. In the model with no interaction term, the effect sizes for PVs and MD measures were not materially affected by mutual adjustment (Table S11). However, when including an interaction term, we observed some evidence of a negative interaction between the overall burden of PVs and PD for breast cancer risk (OR_int_=0.79; 95%CI_int_=0.62,1.00; P_LRT_=0.047, Table S12). This appeared to be largely driven by a positive interaction between NDA and PV burden (OR_int_=1.28; 95%CI_int_=0.97,1.71). For individual genes, the interaction was nominally significant for *ATM* and PD and DA (OR_int_=0.52; 95%CI_int_=0.26,0.99 and 0.50; 95%CI_int_=0.24,0.98 respectively), *CHEK2* and NDA (OR_int_=1.49; 95%CI_int_=1.01,2.23) and *PALB2* and DA (OR_int_=3.70; 95%CI_int_=1.22,16.67). After synthesising sequencing and array genotyping data, the evidence of interaction was stronger for *CHEK2* c.1100delC and NDA (OR_int_=1.42; 95%CI_int_=1.04,1.95, Table S13).

The joint effects were also examined by considering the associations between MD and breast cancer among PV carriers. For example, the ORs for the association between PD and breast cancer were 1.47 (95%CI=1.41,1.54) in the overall population and 1.19 (95%CI=0.93,1.52) in PV carriers (Table S14). The weaker effect size was more apparent for PVs in the moderate-risk genes (OR=1.14; 95%CI=0.84,1.56) than in the high-risk genes (OR=1.41; 95%CI=0.91,2.22).

### MR analysis

We conducted MR analyses based on genetic instruments for MD derived from the SNP associations identified by Sieh(2020) ^4^ and Chen(2022) ^7^ (Table S15). There was evidence for an association between genetically predicted MD measures and breast cancer risk (Figure 3, Table S16, Figure S1-6). The ORs per standard deviation from weighted-median MR for the association between PD and overall breast cancer were 1.54 (95%CI=1.31,1.80) for Sieh(2020) and 1.74 (95%CI=1.49,2.03) for Chen(2022). In addition, associations were seen for all subtypes but were somewhat weaker for triple-negative breast cancer (TNBC): for PD, the ORs were 1.28 (95%CI=0.99,1.66) and 1.48 (95%CI=1.16,1.88) using the Sieh and Chen data, respectively.

**Figure 3.**
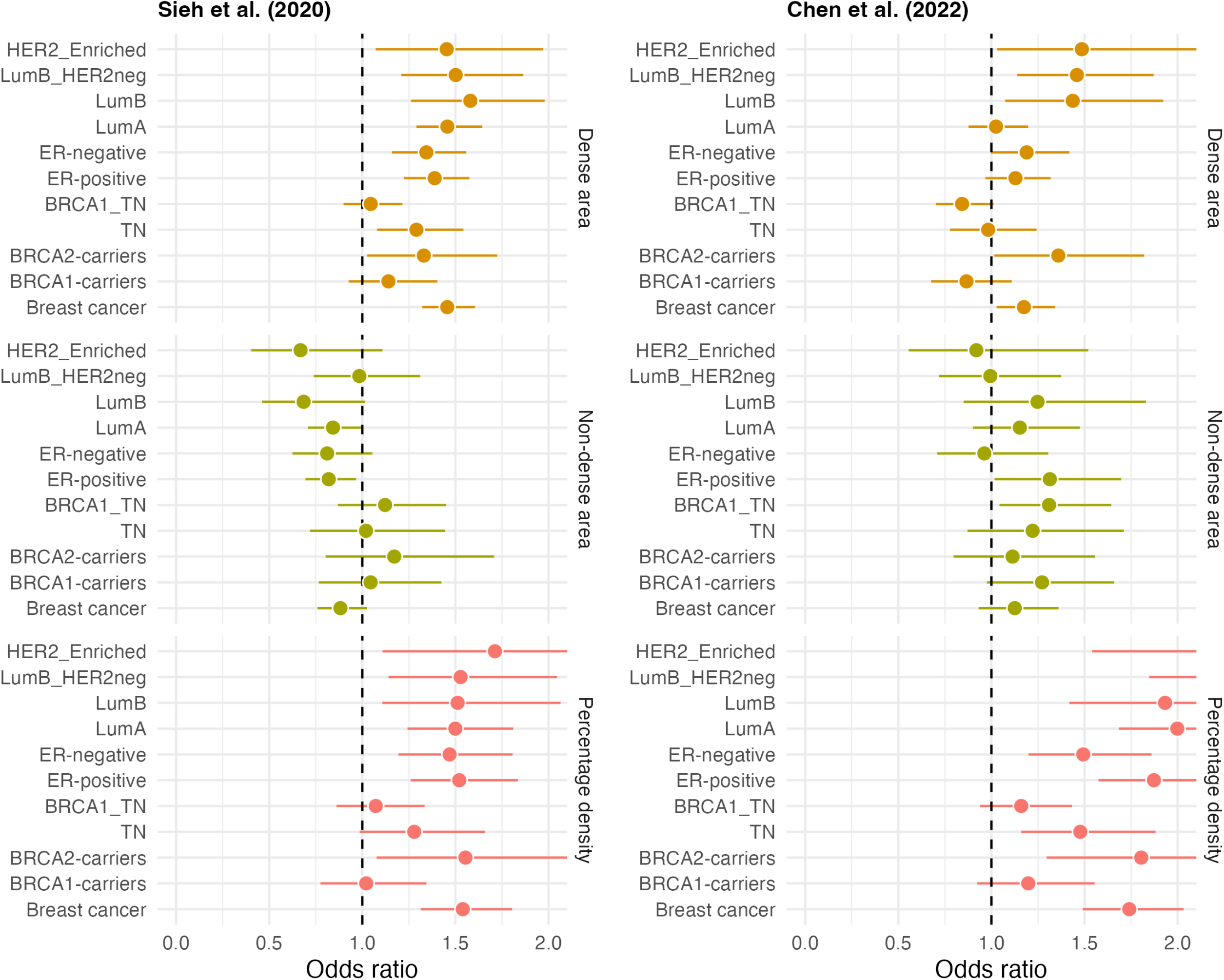
Forest plot of Mendelian randomisation results by using genetic instruments from Sieh et al. (2020) and Chen et al. (2022) (weighted-median method). The x-axis is the estimated odds ratio and 95% confidence interval per one standard deviation change of mammographic density, TN: triple-negative breast cancer, LumA: Luminal-A breast cancer, LumB: Luminal-B breast cancer, LumB_HER2: Luminal B/HER2-negative-like, HER2_Enriched: HER2-enriched-like breast cancer, ERpos: ER-positive breast cancer, ERneg: ER-negative breast cancer, BRCA1_TN: meta-analysis of GWAS of breast cancer in *BRCA1 mutation ca*rriers and GWAS of triple-negative breast cancer, *BRCA1*-carriers: breast cancer in *BRCA1 carriers, BRCA2*-carriers: breast cancer in *BRCA2* carriers.

The associations between genetically predicted MD measures and breast cancer risk in *BRCA1* PV carriers, using summary statistics from CIMBA, were significantly attenuated compared to the overall breast cancer effect sizes. The ORs for genetically predicted PD and breast cancer in *BRCA1* PV carriers were 1.02 (95%CI=0.78,1.34) using Sieh(2020) and 1.20 (95%CI=0.92,1.55) using Chen(2022). For *BRCA2* PV carriers, the effect sizes were similar to those for overall breast cancer (OR=1.55; 95%CI=1.08,2.24, P=0.019 and OR=1.80; 95%CI=1.30,2.51, P=4.65×10^-4^).

The intercepts from the MR-Egger analyses indicated no evidence of pleiotropy, but the Q-statistics indicated significant heterogeneity among the effect estimates. Re-estimation of associations after removing outlier SNPs identified yielded similar effect estimates, both in the overall breast cancer, by subtype and in *BRCA1* and *BRCA2* carriers (Table S16, Figure S1-6).

## Discussion

To our knowledge, this is the first large study to investigate MD in combination with PVs in established breast cancer susceptibility genes, tested systematically, and association with breast cancer. Previous studies that have examined the effect of MD on breast cancer risk among *BRCA1* and *BRCA2* PV carriers have yielded conflicting results. ^9-11^ The sample size in previous studies was, however, small and moreover, these studies were in women identified as carriers through clinical testing, leading to potential confounding with family history.

MD measures and PVs in breast cancer susceptibility genes are both major risk factors for breast cancer and are considered in validated risk models such as BOADICEA. ^25, 26^ Determining the joint effects of these risk factors is, therefore, important for providing valid risk predictions. Consistent with previous studies by BCAC and others, ^1, 27, 28^ we found that both MD measures and the burden of PVs in risk genes were strongly associated with breast cancer risk. We found little evidence of association between the presence of PVs and any of the MD measures, either for all genes combined or any specific gene. Analyses including both MD measures and PVs in the same model made little difference to the effect sizes of either measure, confirming that MD and gene variants are independent risk factors, which is consistent with assumptions in the BOADICEA model.

We did, however, find some evidence of a negative interaction between PD and the burden of PVs, both in aggregate and for individual genes. The interaction was only nominally significant for *ATM*, but the point estimate was less than one for five of the seven genes, including *BRCA1, BRCA2* and *CHEK2*. The association appeared to be largely driven by a positive interaction with NDA.

NDA is negatively associated with breast cancer risk, so the positive interaction is consistent with a weaker association among PV carriers. For example, NDA was associated with an estimated interaction OR of 1.46 for *CHEK2*, which would be consistent with no association in *CHEK2* PV carriers. Similarly, an interaction of OR of 0.77 for PD and PVs in all genes would be consistent with a weaker association with MD in PV carriers. We note, however, that the confidence intervals on these risk estimates were wide. Moreover, *CHEK2* PVs are known to be associated with other traits, including later menopause, and it is possible the interaction seen between MD measures and breast cancer risk might be related to residual confounding by other breast cancer risk factors.

The MR analyses provide a complementary approach to investigating the association between MD measures and breast cancer risk. MR analyses are advantageous for studying causality as they are not subject to biases inherent in observational studies. The MR methodology relies on some key assumptions, characterised as “relevance”, “independence”, and “exclusion restriction”.^29, 30^ The “relevance” assumption is clearly met, while the “independence” assumption is likely to be reasonable given that the GWAS analysis was conducted in European ancestry populations with adjustment for population structure by principal components. The “exclusion restriction” assumption is difficult to verify, and it is possible that variants in genes associated with MD are also associated with breast cancer independently. The MR-Egger analyses, however, did not indicate any evidence for horizontal pleiotropy.

In these MR analyses, there was clear evidence of associations between genetically predicted MD measures and breast cancer risk, with a relative risk per one SD of ∼1.4-1.6 for PD, depending on the SNP set included. This effect size is consistent with that observed in observation studies, including the present study, providing some reassurance on the validity of the MR approach. However, there was no evidence of a comparable association in *BRCA1* PV carriers. The confidence limits in *BRCA1* PV carriers were wide but excluded the overall breast cancer estimate. For *BRCA2* PV carriers, we observed effects comparable to those for overall breast cancer, though the confidence limits were also wide.

There is evidence that individuals with denser breasts exhibit higher expression of DNA damage signals and altered DNA response in contrast to those with lower-density breast tissues. ^31^ The breast cancer susceptibility genes considered here are all involved in DNA repair. ^32, 33^ Therefore, it is plausible, *a priori*, that PVs in these breast cancer susceptibility genes could be related to MD, but we found no evidence for this.

The attenuation of the relative risk for breast cancer associated with MD in *BRCA1* (but not *BRCA2*) PV carriers seen in the MR analyses may be attributable to subtype-specificity: *BRCA1* PVs more strongly predisposed to TNBC. Previous observational studies have shown that MD is a risk factor for all “intrinsic” subtypes of breast cancer, though there is some evidence of a weaker association for TNBC. ^34^ Several case-only studies reported a negative association between MD and TNBC compared to Luminal-A or other types of breast cancer.^35-37^ Interestingly, the MR analyses showed weaker (non-significant) associations for TNBC than the other subtypes. A related point is that breast cancer cases among *BRCA1* PV carriers occur at a younger age: while there is no clear evidence that the association of MD with breast cancer varies with age,^34, 38^ the data at younger ages are more limited. These explanations would not, however, explain an attenuation of the effect in carriers of PVs in moderate-risk genes such as *ATM* and *CHEK2*, which are predominately associated with ER-positive disease. This attenuation, if substantiated, could reflect a modified mechanistic role of MD on breast cancer development in individuals with defective DNA repair.

This study has several important strengths, in particular, the use of a large dataset with MD scored by well-validated quantitative methods and genetic data generated in a single experiment with consistent quality control. We were able to conduct sensitivity analyses and adjust for the main potential confounders. The complementary MR analyses supplied more powerful and less biased results and were qualitatively consistent with observation studies.

We combined data on MD from two distinct methods, which are highly correlated and similarly associated with breast cancer risk. ^39^ Analyses showed no important differences by scoring method. One limitation of our analyses was that they were largely confined to individuals of European ancestry. Given evidence suggesting that the distribution of MD is different in women of Asian ancestry,^40, 41^ further analyses in Asian and other populations will be necessary to confirm the generalisability of these findings. Another limitation was the relatively small number of common genetic variants associated with MD measures. Larger MD GWAS would provide more powerful genetic instruments for MR analyses. Finally, recent studies utilising machine learning approaches applied to large datasets of mammographic images have developed alternative scores that are more predictive than MD alone for both short- and long-term breast cancer risk. ^27, 42, 43^ It will be important to establish whether such measures are also predictive of risk in PV carriers.

## Conclusions

Our results indicate that PVs in breast cancer susceptibility genes are not associated with MD measures. The association between MD and breast cancer risk may be weaker in PV carriers, possibly driven by a weaker association for TNBC. Evaluation in other large datasets will be required to obtain more precise estimates of breast cancer risks associated with MD in PV carriers.

## Data Availability

Requests for individual-level data used in these analyses should be made via the BCAC data access coordinating committee. Summary-level data from BCAC and CIMBA used in the Mendelian Randomisation analysis is publicly available at https://www.ccge.medschl.cam.ac.uk/breast-cancer-association-consortium-bcac), https://www.ccge.medschl.cam.ac.uk/consortium-investigators-modifiers-brca12-cimba and the GWAS Catalog (https://www.ebi.ac.uk/gwas/)

https://www.ccge.medschl.cam.ac.uk/consortium-investigators-modifiers-brca12-cimba

## Funding and Acknowledgements

This work was supported by Cancer Research UK grant: PPRPGM-Nov20\100002 and by core funding from the NIHR Cambridge Biomedical Research Centre (NIHR203312) [*]. *The views expressed are those of the author(s) and not necessarily those of the NIHR or the Department of Health and Social Care. Additional funding for BCAC is provided by the Confluence project which is funded with intramural funds from the National Cancer Institute Intramural Research Program, National Institutes of Health, the European Union’s Horizon 2020 Research and Innovation Programme (grant numbers 634935 and 633784 for BRIDGES and B-CAST respectively), and the PERSPECTIVE I&I project, funded by the Government of Canada through Genome Canada and the Canadian Institutes of Health Research, the Ministère de l’Économie et de l’Innovation du Québec through Genome Québec, the Quebec Breast Cancer Foundation. The EU Horizon 2020 Research and Innovation Programme funding source had no role in study design, data collection, data analysis, data interpretation or writing of the report.

## Author contributions

Prof Easton had full access to all of the data in the study and took responsibility for the integrity of the data and the accuracy of the data analysis.

Concept and design: Easton Statistical analysis: Zhang, Eriksson, Mavaddat, Dennis Acquisition, analysis, or interpretation of data and critical revision of the manuscript for important intellectual content: All authors.

Manuscript writing: Zhang, Easton Administrative, technical, or material support: Lush, Wang, Naven, Bolla

